# Evaluating the effect and mechanism of Yiqi Huayu Jiedu decoction combined with FLOT regimen neoadjuvant chemotherapy for the patients with locally advanced gastric cancer: protocol for a prospective, double-arm, randomized controlled clinical trial

**DOI:** 10.1101/2024.01.24.24301708

**Authors:** Kun Zou, Pei-Chan Zhang, Chun-Yang Luo, Rui Wang, Shuo Xu, Chun-Jie Xiang, Xiang-Kun Huan, Wen-Chao Yao, Xiu-Yuan Li, Jun-Feng Zhang, Shen-Lin Liu, Zhen-Feng Wu

**Author notes:** **Corresponding author and email address:** Zhen-Feng Wu, MD, Department of Surgical Oncology, Jiangsu Province Hospital of Chinese Medicine, Affiliated Hospital of Nanjing University of Chinese Medicine, No. 155 Hanzhong Road, Nanjing 210029, People’s Republic of China. These authors contributed equally to this study (Kun Zou, Pei-Chan Zhang, Chun-Yang Luo).

## Abstract

**Background:** Neoadjuvant chemotherapy plays a vital role in the treatment of advanced gastric cancer, however, optimizing its effectiveness remains an important research focus. Traditional Chinese medicine (TCM), a promising adjunctive therapy, has shown enhanced clinical outcomes when combined with postoperative adjuvant chemotherapy. Therefore, this study is designed to evaluate the clinical efficacy of Yiqi Huayu Jiedu decoction combined with neoadjuvant chemotherapy FLOT in the treatment of advanced gastric cancer.

**Methods:** This study is a prospective, double-arm, randomized controlled trial. It involves a total of 260 patients diagnosed with advanced gastric cancer, who will be randomly assigned to two groups - a TCM treatment group and a control group, each comprising 130 patients. All patients will receive standard FLOT chemotherapy, and patients in the TCM treatment group will additionally receive traditional Chinese medicine treatment with Yiqi Huayu Jiedu decoction. After four cycles of chemotherapy, gastric cancer D2 radical surgery will be performed. The primary objective is to evaluate the postoperative pathological response rate of the tumor. The secondary objectives include evaluating the perioperative nutritional status, the efficacy of traditional Chinese medicine syndrome, and adverse events associated with both chemotherapy and surgery.

**Discussion:** Currently, no trials have investigated the impact of traditional Chinese medicine in combination with neoadjuvant chemotherapy on the preoperative treatment in patients with advanced gastric cancer.

Accordingly, it is imperative to conduct this prospective study to evaluate the clinical efficacy and safety of this regimen, meanwhile providing high-level clinical evidence for traditional Chinese medicine combined with neoadjuvant chemotherapy and introducing an innovative regimen for preoperative comprehensive treatment of gastric cancer.

**Trial registration:** ChiCTR2300072742

## Background

Gastric cancer (GC) is the fifth most prevalent cancer and the third leading cause of cancer mortality worldwide[1]. In China, gastric cancer ranks as the second most prevalent among all malignant tumors, in both incidence and mortality rates[2]. Neoadjuvant chemotherapy has been included in the clinical guidelines for the treatment of gastric cancer, and a large number of previous studies have demonstrated its efficacy and safety [3, 4]. Notably, with the publication of the FLOT4-AIO study results[5, 6], the FLOT regimen, combining 5-fluorouracil, tetrahydrofolate, oxaliplatin and docetaxel, showed a higher pathological response rate and an overall survival outcome, thus it has been considered as a standard neoadjuvant chemotherapy regimen for locally advanced resectable gastric or gastro-oesophageal junction (GEJ) adenocarcinoma. Consequently, the study on neoadjuvant FLOT chemotherapy regimen in gastric cancer has been widely concerned around the world. The results of a real-world data study in Italian confirm the feasibility of FLOT in an unselected population, representative of the clinical practice[7]. A phase II study “GASPAR” trial[8], with the participation of 13 centers in France, are currently investigating the safety and efficacy of FLOT combined with neoadjuvant immunotherapy in the treatment of gastric cancer. Meanwhile, the phase III “PREVENT”-(FLOT9) trial of the AIO /CAOGI /ACOFLOT9 will explore the clinical efficacy of FLOT combined with Hyperthermic IntraPEritoneal Chemoperfusion (HIPEC)[9].

However, at present, there is no relevant research report on the combination of FLOT regimen and traditional Chinese medicine (TCM) in the treatment of gastric cancer. TCM has become an indispensable part of the comprehensive treatment of gastric cancer[10, 11]. Yiqi Huayu Jiedu decoction (YHJD) established by Professor Shenlin Liu, the national famous Chinese Physician in Affiliated Hospital of Nanjing University of Chinese Medicine, has been widely used in clinical practice. Our previous multi-center clinical study [12] has confirmed that YHJD decoction is effective in improving DFS rate in patients with gastric cancer stage III after radical gastrectomy. Meanwhile, another multi-center clinical study on the efficacy of YHJD for reducing the risk of postoperative recurrence and metastasis is being conducted [13]. However, these clinical studies are all combined with postoperative adjuvant chemotherapy, and no study was reported to evaluate whether the combination of YHJD and neoadjuvant chemotherapy can improve the postoperative pathological response rate of advanced gastric cancer.

Accordingly, the present randomized controlled study is designed with the intent to furnish robust clinical evidence for the combined treatment of YHJD alongside the neoadjuvant chemotherapy FLOT regimen in advanced gastric cancer and to provide a novel application scheme for the treatment of traditional Chinese medicine in gastric cancer.

## Methods/design

This study is a prospective, double-arm, randomized controlled trial conducted by Jiangsu Provincial Hospital of Traditional Chinese Medicine. A total of 260 gastric cancer patients will be randomly divided into a TCM treatment group and a control group in a 1:1 ratio, with 130 patients in each group. All enrolled patients have been pathologically confirmed to have gastric adenocarcinoma, and their clinical staging is determined based on the imaging results, following the 8th edition of the UICC clinical TNM staging for gastric cancer (cTNM), diagnosed as cT2-4bN0-3M0. All patients will receive standard FLOT chemotherapy, and patients in the TCM treatment group will additionally receive traditional Chinese medicine treatment with Yiqi Huayu Jiedu decoction. After four cycles of chemotherapy, gastric cancer D2 radical surgery will be performed. Postoperative pathology will be assessed according to the Becher standard for tumor regression staging. The study protocol is shown in **Figure 1**.

**Figure 1.**
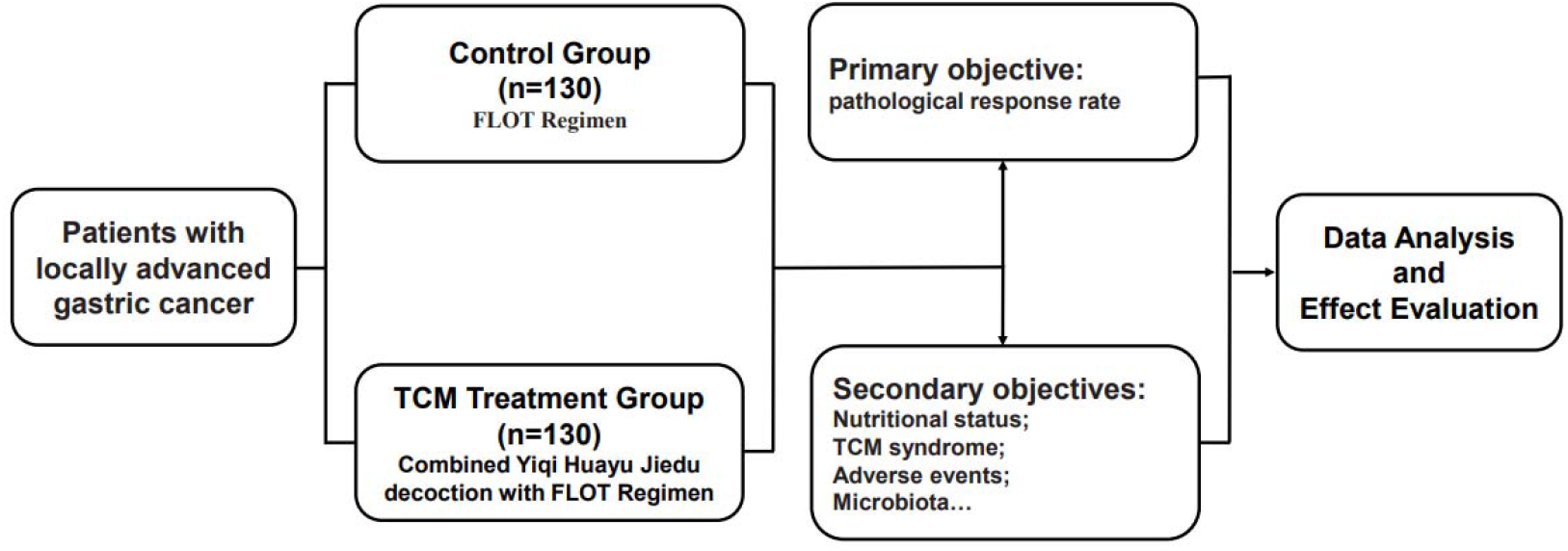
The research flowchart.

This study has been approved by the Ethics Committee of Jiangsu Provincial Hospital of Traditional Chinese Medicine (Ethical approval number: 2023NL-046-02) and registered with the Chinese Clinical Trial Registry (Registration number: ChiCTR2300072742, protocol version number: V4.0, 25 June 2023). All items from the World Health Organization Trial Registration Data Set can be queried via the following website (https://trialsearch.who.int/Trial2.aspx?TrialID=ChiCTR2300072742).

### Patient eligibility and statistical design

#### 1. Diagnostic criteria for locally advanced gastric cancer

According to the “Chinese Society of Clinical Oncology (CSCO) Gastric Cancer Diagnosis and Treatment Guidelines 2022”, endoscopic and imaging examinations are used for the qualitative diagnosis and localization of gastric cancer. The definitive diagnosis of gastric cancer relies on histopathological examination. The diagnosis is gastric adenocarcinoma or adenocarcinoma at the gastroesophageal junction. Staging follows the 8th edition of the UICC Gastric Cancer Clinical TNM staging (cTNM), and patients are diagnosed with cT2-4bN0-3M0 stage.

#### 2. Criteria for grading disease severity

The clinical staging (cTNM) and pathological staging (pTNM) of gastric adenocarcinoma were performed according to the 8th UICC gastric cancer TNM staging system.

#### 3. Sample Size Calculation

According to literature data [5], the postoperative pathological remission rate for the standard FLOT regimen is 37%. Based on clinical observations, it is estimated that the postoperative pathological remission rate for the combination of Yiqi Huayu Jiedu decoction with the FLOT regimen is approximately 55%. Assuming a one-sided α of 5% and a power of 1-β of 80%, a sample size of 117 cases is required. Accounting for a 10% dropout rate, the calculated total is 130 cases. The calculation formula is as follows:

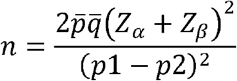

#### 4. Randomization

Randomization was performed using SPSS software. The randomization sequence was generated by the statistical lead of the research group, who had received GCP training. Codes were generated using the Random Number Generator feature in SPSS. A random sequence was assigned in a 1:1 ratio to 260 cases, with 130 cases in each group. Random numbers were placed in sealed envelopes, and each envelope was assigned a serial number based on the order in which the random numbers were generated. Envelopes were then distributed to subjects as they were enrolled. Patients were assigned via telephone contact or text message after meeting the inclusion criteria and providing informed consent.

#### 5. Choice of comparators

The control group received the FLOT regimen as neoadjuvant chemotherapy, while the treatment group received the Yiqi Huayu Jiedu Decoction in combination with the FLOT regimen. The FLOT regimen was chosen as the control based on established principles of efficacy, safety, and comparability. Patients in the control group underwent four cycles of standard FLOT chemotherapy. This regimen has been validated as safe and effective in large randomized controlled trials. Additionally, it is a Grade I recommended regimen for Stage II (cT1-2N1-3M0, cT3-4N0M0) and Stage III (cT3-4aN1-3M0) according to the “Chinese Society of Clinical Oncology (CSCO) Gastric Cancer Diagnosis and Treatment Guidelines 2022.”

### Eligibility criteria

Inclusion criteria and Exclusion criteria are precised in Table 1.

**Table 1.** Eligibility criteria.

| Inclusion criteria | Non-inclusion criteria |
| --- | --- |
| •Patients who meet the diagnostic criteria for locally advanced gastric cancer and are diagnosed with Spleen and Stomach Qi Deficiency with Blood Stasis Obstruction according to Traditional Chinese Medicine (TCM) syndrome differentiation, and have not received relevant treatment in the past.<br>•Patients aged between 18 and 80 years, of any gender.<br>•According to the Eastern Cooperative Oncology Group (ECOG) performance status, patients should have a physical status ranging from 0 to 2. Admission assessments should meet the following criteria: hematopoietic function (white blood cell count $> 3.0 \times 10^9/L$, platelet count $> 100 \times 10^9/L$); liver function (total bilirubin $\leq 1.5 \times ULN$, alanine transaminase $\leq 3.5 \times ULN$, aspartate transaminase $\leq 3.5 \times ULN$); cardiac function (assessed by echocardiography with an ejection fraction $> 50\%$); renal function (serum creatinine $\leq 1.5 \times ULN$, or calculated glomerular filtration rate $> 50 \text{ mL/min}$). There should be no severe complications that would lead to an expected survival of less than 5 years.<br>•Patients must provide informed consent, be willing to participate in the study, and commit to following the protocol. The process of obtaining informed consent should comply with Good Clinical Practice (GCP) regulations. | •Patients who have been diagnosed with or highly suspected of having distant metastases, retroperitoneal lymph node metastases, unresectable tumors with local invasion, recurrent gastric cancer, secondary malignancies, or a history of chemotherapy or radiation therapy will be excluded.<br>•Patients with acute infectious diseases, uncontrolled systemic diseases or complications, or severe primary heart, liver, lung, kidney, blood, or lifethreatening illnesses, known contraindications, or allergies to chemotherapy drugs or traditional Chinese medicine will be excluded.<br>•A history of prior or concurrent malignant tumors, a clear tendency for gastrointestinal bleeding and/or coagulation disorders, poorly controlled blood pressure, and a history of myocardial infarction within the past 6 months will be exclusion criteria.<br>•Prior use of other treatments for advanced gastric cancer, whether traditional Chinese or Western medicine, after the onset of the current illness.<br>•Pregnant or lactating women.<br>•Individuals with intellectual or behavioral impairments that prevent them from providing informed consent.<br>•Suspected or confirmed history of alcohol or substance abuse.<br>•Failure to meet inclusion criteria in terms of various indicators on admission, such as liver, kidney, heart, bone marrow hematopoiesis, and other functions.<br>•Other conditions, as judged by the investigator, that may reduce the likelihood of enrollment or complicate enrollment, such as frequently changing work environments that may lead to loss to follow-up.<br>•Participation in other research projects within the last month. |

### Study assessments

The overview of study assessments and procedures is detailed in Table 2.

**Table 2.** Assessment and intervention schedule for the trial.

|  | Before<br>inclusion | During treatment (±2days) |  |  |  | Before<br>surgery<br>(±7days) | After<br>surgery<br>(±7days) |
| --- | --- | --- | --- | --- | --- | --- | --- |
|  | D0 | D1 | D15 | D29 | D43 | D64 | D78 |
| Diagnosis and eligibility criteria | √ |  |  |  |  |  |  |
| Signed informed consent | √ |  |  |  |  |  |  |
| Randomization information | √ |  |  |  |  |  |  |
| Baseline characteristics | √ |  |  |  |  |  |  |
| **Clinical assessment** |  |  |  |  |  |  |  |
| Pathological response |  |  |  |  |  |  | √ |
| perioperative nutritional status |  | √ | √ | √ | √ | √ | √ |
| Traditional Chinese Medicine syndrome |  | √ | √ | √ | √ | √ | √ |
| **Biological assessment** |  |  |  |  |  |  |  |
| Microbiota indicators | √ |  |  |  |  | √ |  |
| Microbiota-related metabolic markers | √ |  |  |  |  | √ |  |
| EMT |  |  |  |  |  |  | √ |
| Immune cell-related indicators |  |  |  |  |  |  | √ |
| **Safety assessment** |  |  |  |  |  |  |  |
| Chemotherapy-related adverse events |  | √ | √ | √ | √ | √ |  |
| Surgery-related adverse events |  |  |  |  |  |  | √ |

#### Primary objective

The main efficacy evaluation criterion for assessing the clinical effectiveness of combining the Yiqi Huayu Jiedu decoction with the FLOT regimen in the treatment of locally advanced gastric cancer was the postoperative pathological response rate.

#### Secondary objectives

Additionally, the research assessed the effectiveness, safety, and quality of life of the FLOT regimen when combined with the Yiqi Huayu Jiedu decoction. The study encompassed evaluations of perioperative nutritional status, the efficacy of Traditional Chinese Medicine syndrome, and adverse events associated with chemotherapy and surgery. Furthermore, microbiota indicators, microbiota-related metabolic markers (short chain fatty acid), markers associated with EMT, and immune cell-related indicators were examined. The objective was to assess the structural changes and significance of gastrointestinal flora in response to the treatment involving the Yiqi Huayu Jiedu decoction combined with neoadjuvant chemotherapy. Moreover, the study aimed to explore the potential mechanism underlying this treatment, particularly how gastrointestinal flora might participate in enhancing the tumor-suppressing functions of specific immune cells in the tumor microenvironment.

### Treatment

This study is divided into two regimens: the FLOT regimen and Yiqi Huayu Jiedu decoction.

#### 1. FLOT Regimen Medications

Docetaxel, specification: 20mg; Oxaliplatin, specification: 50mg; 5-Fluorouracil, specification: 250mg; Leucovorin, specification: 100mg. These medications are selected from the National Drug Procurement List and have been evaluated for therapeutic equivalence.

#### 2. Yiqi Huayu Jiedu decoction

Astragalus root (Huang Qi): 45g; Codonopsis Pilosula (Dang Shen): 15g; Rhizoma Sparganii (San Leng): 20g; Curcuma Zedoary (E Zhu): 20g; Rhizoma Atractylodis Macrocephalae (Chao Bai Zhu): 10g; Radix Paeoniae Alba (Chao Bai Shao): 10g; Angelica Sinensis (Dang Gui): 10g; Pericarpium Citri Reticulatae (Chen Pi): 10g; Salvia Chinensis (Shi Jian Chuan): 30g; Oldenlandia diffusa (Baihuasheshecao): 30g. The herbs for this formula are provided and decocted by Jiangsu Provincial Hospital of Traditional Chinese Medicine.

### Method of Administration

#### 1. FLOT Regimen

A standard FLOT regimen chemotherapy cycle includes: Intravenous infusion of 5-Fluorouracil (5-FU) at a dose of 2600mg/m2 over 24 hours through a peripherally inserted central catheter (PICC) on day one. Intravenous injection of Docetaxel at a dose of 50mg/m2. Intravenous injection of Oxaliplatin at a dose of 85mg/m2. Intravenous injection of Leucovorin at a dose of 200mg/m2. This cycle is repeated every 14 days. Patients receive a total of four standard FOLT regimen chemotherapy cycles.

#### 2. Combined Yiqi Huayu Jiedu decoction with FLOT Regimen

In addition to the standard FLOT regimen, patients also receive the Yiqi Huayu Jiedu decoction. This formula is synchronized with the FOLT regimen, with one course lasting 14 days, and a total of four cycles. This formula is prepared as a decoction and taken after breakfast and dinner. The medications and decoction service are provided by the Pharmacy Department of Jiangsu Provincial Hospital of Traditional Chinese Medicine.

Note: Modifications or interruptions to the specified dosages in the protocol are allowed. For instance, in the case of adverse events related to chemotherapy, such as febrile neutropenia, thrombocytopenia, or other hematologic toxicities, the dosages of Docetaxel and Oxaliplatin may be adjusted to 75%. If these events recur, the dosages can be further reduced to 50%. For other non-hematologic toxicities exceeding Grade 2, all drug dosages may be adjusted to 75%, with further reductions to 50% if necessary. The treating physician may also consider dose adjustments for specific toxicities at their discretion. Treatment will be discontinued if unacceptable toxicity occurs, if there is disease progression or death, if requested by the patient, or if the treating physician determines that discontinuation is in the best interest of the patient.

### Termination of trial cases

1. Patients who have serious adverse events, such as toxic effects of chemotherapy that are so life-threatening that the clinical trial should be stopped according to the doctor’s judgment.
2. Disease progression during the course of the illness, or the emergence of other conditions that affect trial observations, such as local tumor progression and distant metastasis during treatment that precludes curative surgery. Patients for whom the clinical trial should be terminated based on the physician’s judgment will be considered as invalid cases.
3. Suffered serious deviation in the implementation of clinical trial scheme, such as poor adherence, difficult to evaluate drug effect.
4. The subjects in the process of clinical trials is not willing to continue the clinical trials, to the competent doctors put forward out of the demander clinical trials.

### Dropout and Handling of Cases

#### 1. Drop-out criteria

Participants who have provided informed consent and have been screened for eligibility but did not complete the prescribed treatment course and observation period as outlined in this protocol will be considered as dropouts.

#### 2. Handling of dropped cases

1. When participants drop out, researchers should make efforts to contact them through home visits, scheduled follow-up appointments, telephone calls, or written correspondence. This is to inquire about the reasons for discontinuation, record the last time medication was taken, and complete any assessment items that can still be obtained.
2. For cases that exit the trial due to allergies or other adverse reactions, or because the treatment is ineffective, researchers should take appropriate treatment measures based on the individual circumstances of the participants.
3. Dropped cases should be properly preserved with relevant trial data, both for record-keeping purposes and for inclusion in the overall dataset for statistical analysis. Dropped cases do not need additional supplementation.

#### 3. Exclusion of Cases

1. Cases that do not meet the inclusion criteria but meet the exclusion criteria.
2. Cases that have never used the investigational drug.
3. Cases with no data available after randomization.

Before performing data statistical analysis, a discussion between the statisticians and the principal investigator should determine whether these cases should be excluded.

##### Statistical analysis

A descriptive statistical analysis was performed for quantitative data. For qualitative data, descriptive statistical analysis should be done by frequency table, percentage, or composition ratio. Comparison before and after treatment will be carried out through paired t-test or paired rank-sum test according to the data distribution type. The differences between groups were compared using t-test or rank-sum test for quantitative data and chi-square or rank-sum test for qualitative data. All statistical calculations were performed by SPSS statistical analysis software. Data should be analyzed with 2-sided test and P<0.05 is considered as statistically significant.

##### Data monitoring committee

An Independent Data Monitoring Committee will be set-up to ensure the protection of patients, to ensure the ethical conduct of the study, to evaluate the benefit/risk ratio of the study and to ensure an independent review of the scientific outcomes during and at completion of the study. The committee will include a biostatistician, a pharmacologist and a medical oncologist.

##### Data management

Every patient needs at least one physician to be responsible for enrolling patients in this trial and arranging patient therapy. Two physicians will collect data and complete the case report form (CRF) at their center. The investigator ensures the accuracy, completeness, and timeliness of the data recorded and of the provision of answers to data queries according to the Clinical Study Agreement. The CRF will be regarded as raw material, and all data in the CRF will be archived electronically on a specific computer. All electronic documents will be confidential, and only the data manager will have the password. Only the project leader will have the right to use the database, and other researchers will not be allowed to use the database unless permitted.

## Discussion

Surgical treatment alone cannot improve the poor prognosis of advanced gastric cancer. Multimodal treatment approaches, with neoadjuvant chemotherapy as a representative, have a significant milestone significance in the treatment of advanced gastric cancer[7, 9, 13].

To date, clinical trials investigating the impact of combining traditional Chinese medicine with neoadjuvant chemotherapy for advanced gastric cancer are lacking. Most previous clinical studies on TCM treatment and tumors focused on the postoperative stage. However, our study firstly combines traditional Chinese medicine treatment with preoperative chemotherapy and aims to evaluate the effect of YHJD decoction on the postoperative pathological response rate of gastric cancer after neoadjuvant chemotherapy. Simultaneously, we will collect tumor tissue samples from gastric cancer patients subsequent to their treatment with TCM and neoadjuvant chemotherapy to explore the changes in the immune cells and intratumoral flora in tumor tissues, which will intuitively demonstrate the immunostimulatory effect of drugs on the tumor microenvironment. These findings will give insights into the effect of TCM combined neoadjuvant chemotherapy on tumor immune microenvironment, and provide a research foundation for subsequent determination of the optimal combination regimen of TCM, chemotherapy, and immunotherapy.

Based on the above, the main objective of this randomized controlled study is to explore the clinical efficacy of the YHJD in combination with the FLOT regimen in neoadjuvant chemotherapy for gastric cancer. Furthermore, we aim to use this as a foundation to develop a treatment protocol suitable for clinical application, thus establishing a solid basis for the integration of TCM and chemotherapy in the clinical guidelines for neoadjuvant chemotherapy in gastric cancer.

### Trial status

This study was registered online in June 2023. The recruitment started in July 2023 and is still ongoing.

## Data Availability

All data produced in the present study are available upon reasonable request to the authors

## Disclosure statement

### Ethics Statement

This study has been approved by the Ethics Committee of Jiangsu Provincial Hospital of Traditional Chinese Medicine (Ethical approval number: 2023NL-046-02) and registered with the Chinese Clinical Trial Registry (Registration number: ChiCTR2300072742).

### Availability of data and materials

The datasets used or analyzed during the current study are available from the corresponding author on reasonable request.

### Roles and responsibilities

WZF designed this study and revised the manuscript; ZK drafted this manuscript; PCZ, CYL, RW and WCY are currently involved in study implementation; XYL and HXK were mainly responsible for chemotherapy; SX and CJX contributed to data analysis and interpretation; ZFW, JFZ and SLL participated in designing and conducting the study. All authors contributed to refinement of the study protocol and approved the final manuscript.

